# Impaired humoral immunity to BQ.1.1 in convalescent and vaccinated patients

**DOI:** 10.1101/2022.12.31.22284088

**Authors:** Felix Dewald, Martin Pirkl, Elvin Ahmadov, Martha Paluschinski, Joachim Kühn, Carina Elsner, Bianca Schulte, Maike Schlotz, Göksu Oral, Jacqueline Knüfer, Michael Bernhard, Mark Michael, Maura Luxenburger, Marcel Andrée, Marc Tim Hennies, Wali Hafezi, Marlin Maybrit Müller, Philipp Kümpers, Joachim Risse, Clemens Kill, Randi Katrin Manegold, Ute von Frantzki, Enrico Richter, Dorian Emmert, Werner O. Monzon-Posadas, Ingo Gräff, Monika Kogej, Antonia Büning, Maximilian Baum, Finn Teipel, Babak Mochtarzadeh, Martin Wolff, Henning Gruell, Veronica Di Cristanziano, Volker Burst, Hendrik Streeck, Ulf Dittmer, Stephan Ludwig, Jörg Timm, Florian Klein

## Abstract

Determining SARS-CoV-2 immunity is critical to assess COVID-19 risk and the need for prevention and mitigation strategies. We measured SARS-CoV-2 Spike/Nucleocapsid seroprevalence and serum neutralizing activity against Wu01, BA.4/5 and BQ.1.1 in 1,411 individuals who received medical treatment in five emergency departments in North Rhine-Westphalia, Germany. We detected Spike-IgG in 95.6%, Nucleocapsid-IgG in 24.0% and neutralization against Wu01, BA.4/5 and BQ.1.1 in 94.4%, 85.0%, and 73.8% of participants, respectively. Neutralization against BA.4/5 and BQ.1.1 was reduced 5.6- and 23.4-fold compared to Wu01. Accuracy of S-IgG detection for determination of neutralizing activity against BQ.1.1 was reduced substantially. Furthermore, we explored previous vaccinations and infections as most important correlates of improved BQ.1.1 neutralization using multivariable and Bayesian network analyses. Given an adherence to COVID-19 vaccination recommendations of only 67.7% of all participants, we highlight the need for improvement of vaccine-uptake to reduce the COVID-19 risk in upcoming infection-waves with immune evasive variants.

## Introduction

Population immunity against SARS-CoV-2 will play a key role in the course of the pandemic and determine morbidity and mortality of COVID-19^1–3^. To date, 3 years after the emergence of SARS-CoV-2, immune evasion presents the most significant challenge to combat COVID-19^4–8^.

At the beginning of 2022, a rapid surge of infections was detected worldwide and driven by the Omicron variant that exhibited substantial immune evasion properties^9–11^. Subsequently, multiple Omicron sub-lineages evolved, including BA.5 that accumulated additional mutations in the Spike protein and became the predominant variant globally in June 2022^12^. To that date, BA.5 demonstrated the strongest immune escape from antibodies induced by either SARS-CoV-2 vaccination or infection as well as from therapeutic monoclonal antibodies^13–17^. However, the continuous evolution of the Omicron variant gave rise to further sub-lineages, including BQ.1 and BQ.1.1 with a relative share of all sequenced variants worldwide of 0.1% in August, but 49.7% in November 2022^18,19^. This increase is likely to be caused by additional immune evasion properties in the BQ.1 and BQ.1.1 subvariants, enabling infections of SARS-CoV-2 vaccinated and convalescent individuals. Indeed, early data on neutralization resistance of BQ.1 and BQ.1.1 show that it is mainly driven by a N460K mutation while the R346T and K444T mutations of BQ.1.1 contribute to a lesser extent^20–23^. Furthermore, in comparison with BA.5, enhanced fusogenicity of BQ.1 and BQ.1.1 could be observed^22,24^.

In this multicenter-study, we determined SARS-CoV-2 Spike (S)-IgG levels, Nucleocapsid (NC)-IgG levels and serum neutralization against Wu01, BA.4/5 and BQ.1.1 in 1,411 patients that received medical treatment at emergency departments of five university hospitals in North Rhine-Westphalia, Germany between August and September 2022. We combined IgG levels and neutralization activity with detailed information on medical history and SARS-CoV-2 immune status of the participants. Finally, we conducted multivariable analysis and Bayesian network analysis to provide a better understanding on factors determining quantity and quality of the antibody response to SARS-CoV-2. Our results inform on SARS-CoV-2 immune status and its predictive factors, which helps to assess COVID-19 risk in highly vulnerable groups.

## Results

### Characteristics of study participants to determine SARS-CoV-2 humoral immunity

To determine SARS-CoV-2 point seroprevalence and neutralization activity, we conducted a multi-center cross-sectional seroprevalence study in five emergency departments of university hospitals (maximum care hospitals) in North Rhine-Westphalia, Germany (**Fig. 1a and b**). During the time of sample collection (August and September 2022), 10,191 patients seeked medical treatment in the emergency departments. Of those patients, 1,411 (13.9%) were enrolled for study participation. There was no significant difference between age and sex distributions of all emergency department patients and the study participants, indicating representativeness of the study sample for the source population (**Supplementary Fig. 1, Supplementary Fig. 2**). We collected serum blood samples from all participants and determined S- and NC-IgG reactivity using chemiluminescence immunoassay (CLIA) and enzyme-linked immunosorbent assay (ELISA). Pseudovirus neutralization assays were performed to determine serum neutralizing activity against SARS-CoV-2 Wu01, BA.4/5, and BQ.1.1 variants (**Fig. 1a**). Information on epidemiological and clinical data as well as information on COVID-19 vaccination status and previous infections were collected in structured interviews and extracted from medical records. Enrolled participants had a median age of 53 years (range 18-98, IQR: 35-69) with an overall balanced sex distribution (48.5% female; 51.3% male; **Fig. 1, Supplementary Table 1**). 64.2% of the participants reported pre-conditions related most frequently to cardiovascular (52.3%) and neoplastic (24.4%) diseases (**Fig. 1d**). 13.6% of the participants reported drug immunosuppression at the time of sample collection (**Fig. 1e**). 94.4% of the participants reported to have received at least one immunization with an EU-approved vaccine (**Fig. 1f**) and 45.7% reported at least one previous SARS-CoV-2 infection, resulting in 50.8% of the participants exposed to at least 4 previous S-antigen contacts from either vaccination or infection (**Fig. 1g**). Thus, when stratified by age, 67.7% of all participants were vaccinated according to German COVID-19 vaccination recommendations^25^ (**Fig. 1f**).

**Fig. 1:**
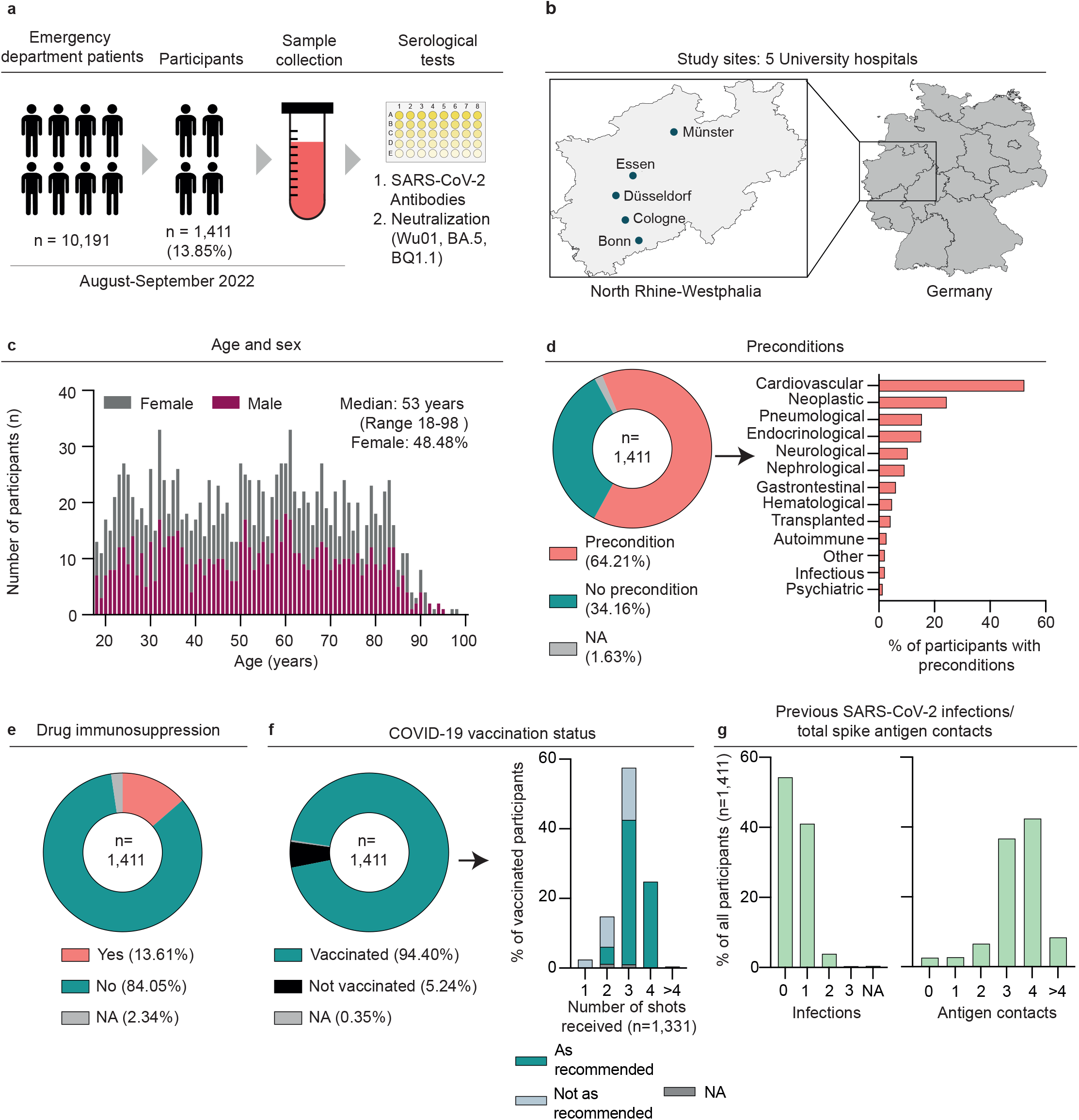
Characteristics of study participants to determine SARS-CoV-2 humoral immunity. (a) Illustration depicting number of study participants recruited in emergency departments, study timeline and experimental procedures. (b) Illustration depicting the locations of the 5 study sites. Maps of Germany and North Rhine-Westphalia were designed with the iMapU tool provided by iExcelU. (c) Distribution of age and sex of the participants. (d) Pie chart depicting presence of pre-conditions. Bar chart illustrating the distribution of pre-conditions stratified by organ-system. (e) Pie chart depicting presence of drug immunosuppression as reported by the participants at the time of sample collection. (f) Pie chart illustrating vaccination status of the participants. Bar chart depicting reported vaccination scheme stratified by number of received shots. Compliance with vaccination recommendations according to age is indicated by corresponding colors. (g) Bar charts illustrating reported previous infections and total Spike antigen contacts consisting of vaccinations and infections.

### High S-IgG sero-prevalence in patients visiting emergency departments in North Rhine-Westphalia, Germany

To determine SARS-CoV-2 humoral immunity, we first measured seroprevalence and levels of S-IgG in all participants. S-IgG could be detected in 95.6% of the participants. Of the 4.4% S-IgG-negative participants, 27.9% reported drug immunosuppression at the time of sampling. Of those reporting no drug immunosuppression, 31.8%, 29.5%, 6.8%, 22.7%, 6.8% and 2.3% reported 0, 1, 2, 3, 4 or > 4 previous S-antigen contacts, respectively (**Fig. 2a**). Thus, considering German COVID-19 vaccination recommendations^25^, 86.9% of the sero-negative participants were either immunosuppressed and/or insufficiently vaccinated. Next, we characterized S-IgG levels stratified by sex, age, pre-conditions, drug immunosuppression, and the number of S-antigen contacts (**Fig. 2b**). S-IgG levels were not significantly different between females and males (GeoMean 1,697 versus 1,836 BAU/ml, Mann-Whitney test: p = 0.663), different age groups (18-30, 31-60, and > 60 years; 2,003, 1,612, and 1,858 BAU/ml; Kruskal-Wallis test: p = 0.357) and participants with or without pre-conditions (GeoMean 1,884 versus 1,698 BAU/ml; Mann-Whitney test: p = 0.538) but lower in participants with drug immunosuppression compared to those without (GeoMean 1,042 versus 1,908 BAU/ml, Mann-Whitney test: p = 0.0005). Furthermore, S-IgG increased with increasing number of reported S-antigen contacts from 86 to 4,450 BAU/ml for 0 to > 4 reported S-antigen contacts (Kruskal-Walis test: p < 0.0001; Dunn’s multiple comparisons tests: 0 versus 1, p > 0.999; 1 versus 2, p = 0.029; 2 versus 3, p = 0.06; 3 versus 4, p < 0.001; 4 versus > 4, p = 0.49). Like the total number of total S-antigen contacts, the number of received vaccinations or previous infections and NC-IgG serostatus affected S-IgG levels significantly (**Supplementary Fig. 3**).

**Fig. 2:**
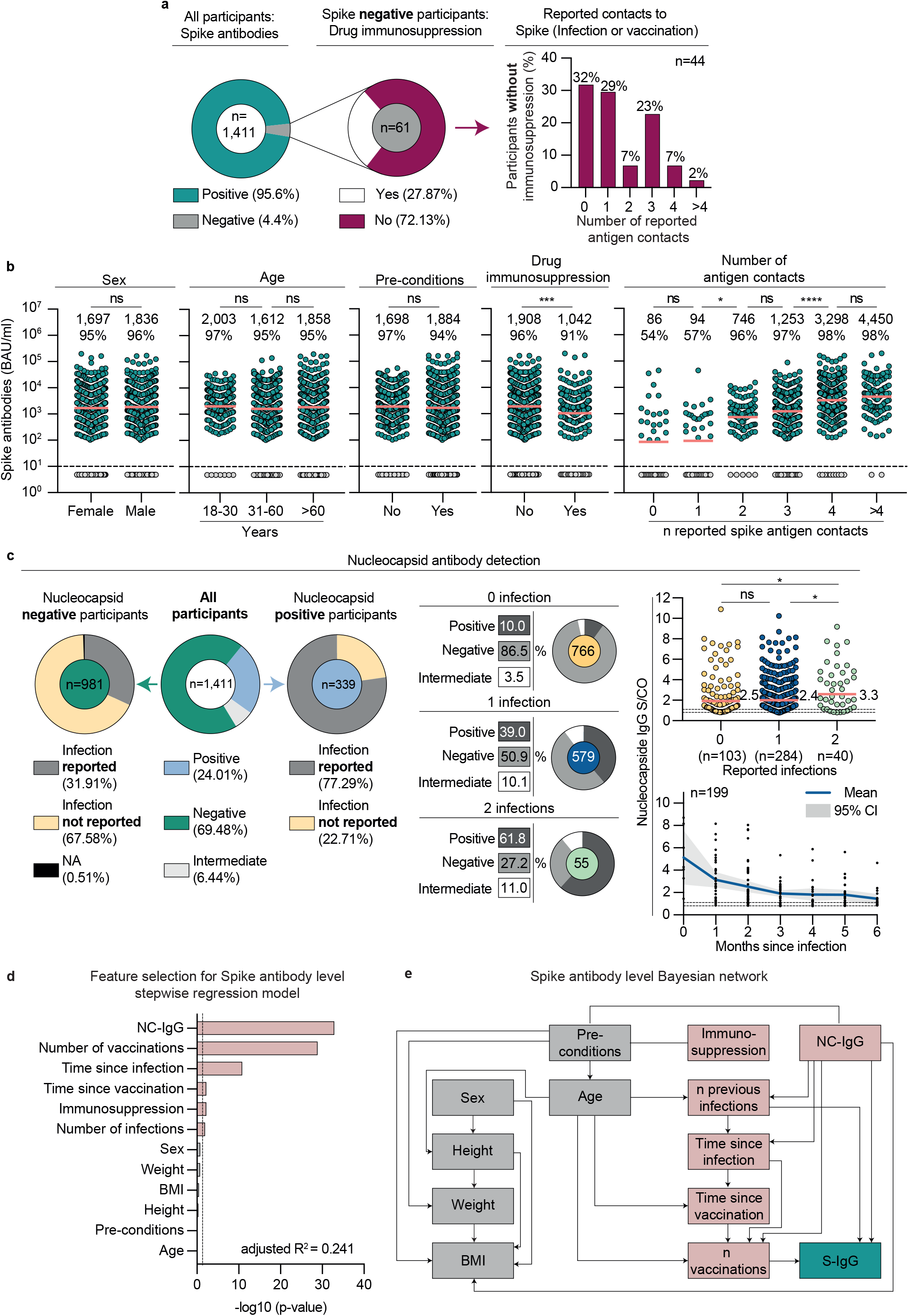
High S-IgG sero-prevalence and factors to predict S-IgG levels. (a) Pie charts illustrating SARS-CoV-2 S-IgG prevalence and presence of drug immunosuppression of Spike negative participants. Bar chart indicating reported number of antigen contacts of participants with no detectable S-IgG and drug immunosuppression. (b) Dot plots depicting S-IgG BAU/ml values, subdivided based on sex, age, pre-conditions, drug immunosuppression, and number of S-antigen contacts. Dotted lines represent the limit of detection (33.8 BAU/ml). Geometric means are indicated by horizontal red lines and listed in each plot over total fractions of participants with detectable S-IgG. Mann-Whitney tests, Kruskal-Wallis-tests and Dunn’s multiple comparisons tests were performed for statistical analyses. Ns, *, **, ***, and **** represent p-values ≥ 0.05, < 0.05, ≤ 0.01, ≤ 0.001, and ≤ 0.0001, respectively. (c) Left pie charts illustrate NC-IgG prevalence and fractions of participants reporting any or reporting no previous infections in NC-IgG positive and negative participants, respectively. Right pie charts depict NC-IgG prevalence stratified by number of reported previous infections. Dot plot illustrates NC-IgG values (S/CO) stratified by number of reported infections. Dotted lines represent cut-off to negative and borderline (0.8 and 1.1 S/CO). Means are indicated by horizontal red lines. Kruskal-Wallis-test and Dunn’s multiple comparisons tests were performed for statistical analyses. * and ns represent p-values < 0.05 and ≥ 0.05, respectively. NC-IgG S/CO dynamic after infection is illustrated stratified by months after infection. Only data of individuals that reported one previous infection are shown. Individual values, mean S/CO and 95% confidence interval are depicted by black dots, blue line, or grey area, respectively. (d) Stepwise forward regression model for predicting S-IgG (BAU/ml) using continuous features (age, height, weight, body mass index (BMI), number of infections, number of vaccinations, time since infection, and time since vaccination) and categorical features (sex, pre-conditions, NC-IgG, and immunosuppression). Features with p < 0.05 in the multivariable regression model are highlighted in red. (e) Bayesian network of the features predicting S-IgG. The graph connects the features, which are predictive of each other with S-IgG as sink. Features with p < 0.05 in the multivariable regression model (D) are highlighted in red.

### NC-IgG seroprevalence contributes to prediction of S-IgG levels

Next, we determined seroprevalence and levels of NC-IgG in all participants. NC-IgG could be detected in 24.0% of the enrolled participants (**Fig. 2c**). Of all NC-IgG-positive participants, 77.3% and of all NC-IgG-negative participants, 31.9% reported at least one previous SARS-CoV-2 infection, respectively. The fractions of NC-IgG-positive participants were higher in those that reported previous infections (10.0%, 39.0%, 61.8% for 0, 1 or 2 previous infections, respectively). Furthermore, NC-IgG values were higher in participants with 2 previous infections compared to those with 1 or no infection (mean S/CO 2.5 and 2.4 for 0 and 1 infection versus 3.3 for 2 infections, Dunn’s multiple comparisons test: 0 versus 1, p = 0.038; 0 versus 2, p = 0.029). In all NC-IgG-positive or -borderline participants with 1 reported infection within the last 6 months (n = 199), mean S/CO decreased from 5.13 to 1.44 during the first 6 months after infection.

Given the observed influence of different clinical and serological features on S-IgG levels, we performed multivariable analysis including sex, height, weight, BMI, age, pre-conditions, drug immunosuppression, number of reported vaccinations and infections, time since last vaccination and infection (months) and NC-IgG serostatus in a stepwise regression model (**Fig. 2d**). Features were only added when they significantly improved the model according to a likelihood ratio test. The resulting model (adjusted R^2^ = 0.241) showed that NC-IgG detection is most predictive for S-IgG levels (p < 2×10^−16^), followed by the number of vaccinations (p < 2×10^−16^), time since infection (p = 0.008), time since last vaccination (p = 0.007), drug immunosuppression (p = 0.007), and number of infections (p = 0.008). Sex, weight, BMI, height, pre-conditions, and age were no significant parameters for S-IgG prediction. Bayesian network analysis revealed that NC-IgG serostatus, number of previous infections and received vaccinations directly predicted S-IgG levels while other parameters were only indirectly predictive (**Fig. 2e**).

### High S-IgG levels correlate with SARS-CoV-2 neutralizing activity

To determine serum neutralization against Wu01 and BA.4/5 variants, we first determined the fraction of participants that showed detectable serum neutralization indicated by serum ID_50_s > 10. Of all participants, 94.4% and 84.9% showed neutralizing activity against Wu01 and BA.4/5, respectively (**Fig. 3a**). The geometric mean of ID_50_ was significantly lower for neutralizing activity against BA.4/5 than against Wu01 (243 vs. 1,440) which corresponds to a 5.92-fold decrease in serum neutralization (Wilcoxon matched-pairs signed rank test: p < 0.0001; **Fig. 3a**). Next, we correlated S-IgG values of all participants with detectable S-IgG against ID_50_ values of all participants with detectable serum neutralization (n=1,176) against Wu01 (r_s_ = 0.74, p < 0.0001) and BA.4/5 (r_s_ = 0.62, p < 0.0001) (**Fig. 3b**). Finally, we computed the fractions of log-tiered serum ID_50_ categories for Wu01 and BA.4/5 stratified by S-IgG levels categorized by log-tiered BAU/ml categories (**Fig. 3c**). No neutralizing activity could be detected against Wu01 and BA.4/5 for S-IgG levels of 10-100 BAU/ml, in 32.4% and 64.7% of the participants, and for S-IgG levels between 100 and 1,000 BAU/ml, in 2.7% and 35.6% of the participants, respectively.

**Fig. 3:**
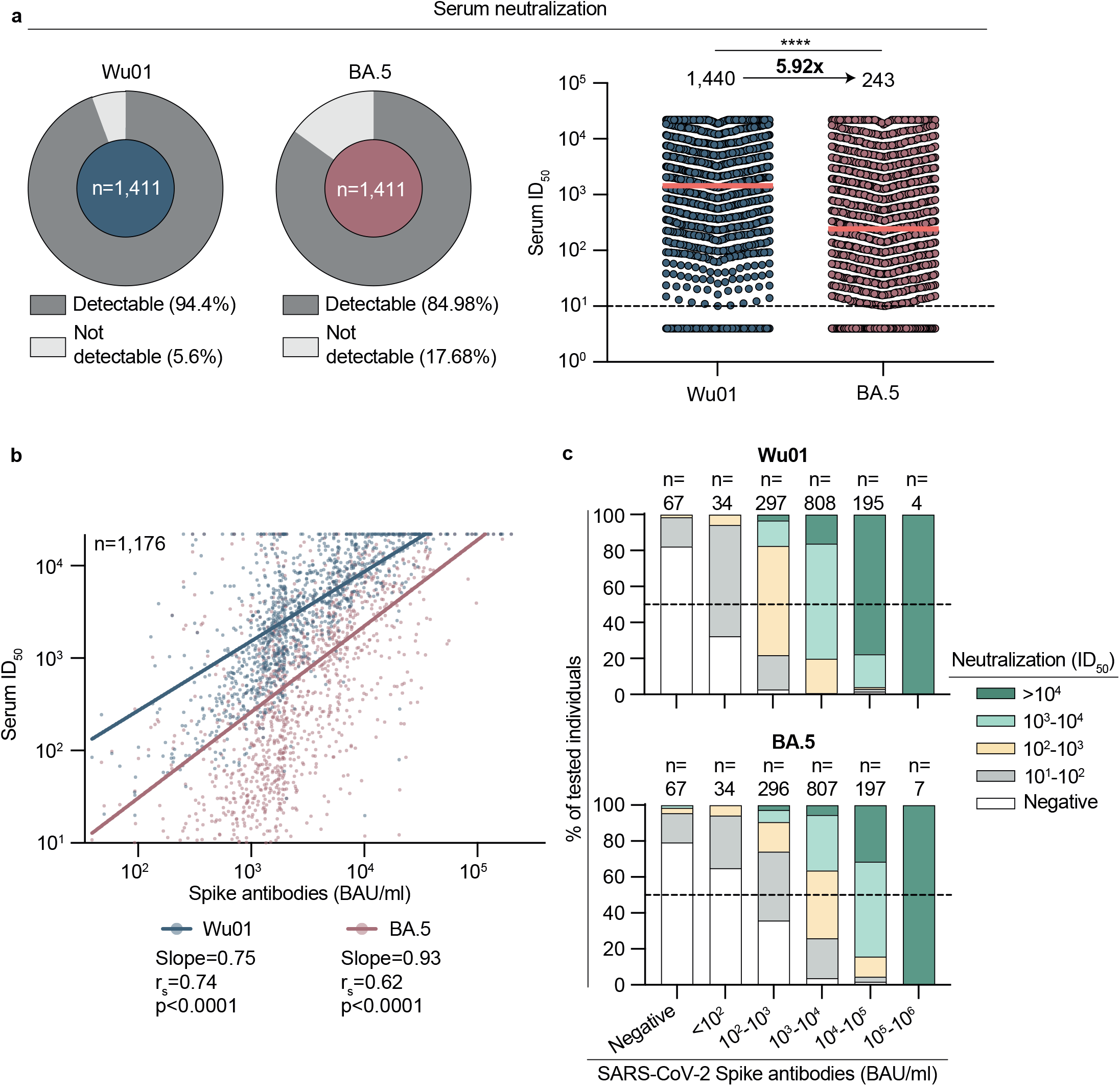
High S-IgG levels correlate with SARS-CoV-2 neutralizing activity. (a) Left: Pie charts illustrating SARS-CoV-2 serum neutralization prevalence (limit of detection ID_50_ = 10). Right: Dot plot depicting serum ID_50_ for Wu01 and BA.4/5 variants. Dotted lines represent the limit of detection (ID_50_ = 10). Geometric means are indicated by horizontal red lines and listed for both variants over total fractions of participants with detectable serum neutralization. Arrow indicates x-fold decrease of serum neutralization. Two-sided Wilcoxon matched pairs signed rank test was performed for statistical analyses. **** represent a p-value < 0.05. (b) Spearman correlation of S-IgG (BAU/ml) against serum neutralization (ID_50_) of Wu01 and BA.4/5 variants indicated by respective colors. Red and blue lines indicate fit lines computed with robust regression. (c) Serum neutralization (ID_50_) is categorized by indicated color and stratified by S-IgG (BAU/ml) for Wu01 und BA.4/5 variants. Dotted lines represent 50% of all participants in each stratum. N of each stratum is indicated on top of the graphs.

We conclude that IgG-detection can assess immunity against SARS-CoV-2, however decreased accuracy for the prediction of serum neutralizing activity introduced by immune evasive variants must be considered.

### S-IgG levels and S-antigen contacts are most predictive for serum neutralization against Wu01 and BA.4/5

To explore predictive features for serum neutralization, we first characterized neutralizing activity against Wu01 and BA.4/5 stratified by sex, age, pre-conditions, drug immunosuppression, number of received vaccinations, number of previous infections, and NC-IgG serostatus (**Fig. 4a**). While serum neutralization was not significantly different between females and males, participants aged 18-30 years had a significantly higher geometric mean of ID_50_ compared to participants aged 31-60 and > 60, both for neutralization against Wu01 and BA.4/5 (ID_50_ GeoMean_Wu01_: 2,306 versus 1,212 and 1,405, Dunn’s multiple comparisons tests: p = 0.002 and p = 0.029; ID_50_ GeoMean_BA.4/5_: 613 versus 231 and 167; Dunn’s multiple comparisons tests: p < 0.0001 and p < 0.0001). Furthermore, serum neutralization was affected by drug immunosuppression at the time of study participation (ID_50_ GeoMean_Wu01_: 1,583 versus 747; ID_50_ GeoMean_BA.4/5:_ 267 versus 117; Dunn’s multiple comparisons tests: p = 0.0006 and p = 0.002) and the presence of pre-conditions (ID_50_ GeoMean_Wu01_: 1,227 versus 1,919; ID_50_ GeoMean_BA.4/5:_ 190 versus 385; Dunn’s multiple comparisons tests: p = 0.004 and p = 0.0001). Serum neutralization against Wu01 and BA.4/5 was significantly higher in vaccinated participants in comparison to unvaccinated (Kruskal-Wallis test: p < 0.0001), in participants that reported previous infections compared to those that did not (Kruskal-Wallis test: p < 0.0001) and in participants with detectable NC-IgG levels compared to those without (Mann-Whitney test: p < 0.0001) (**Fig. 4a**).

**Fig. 4:**
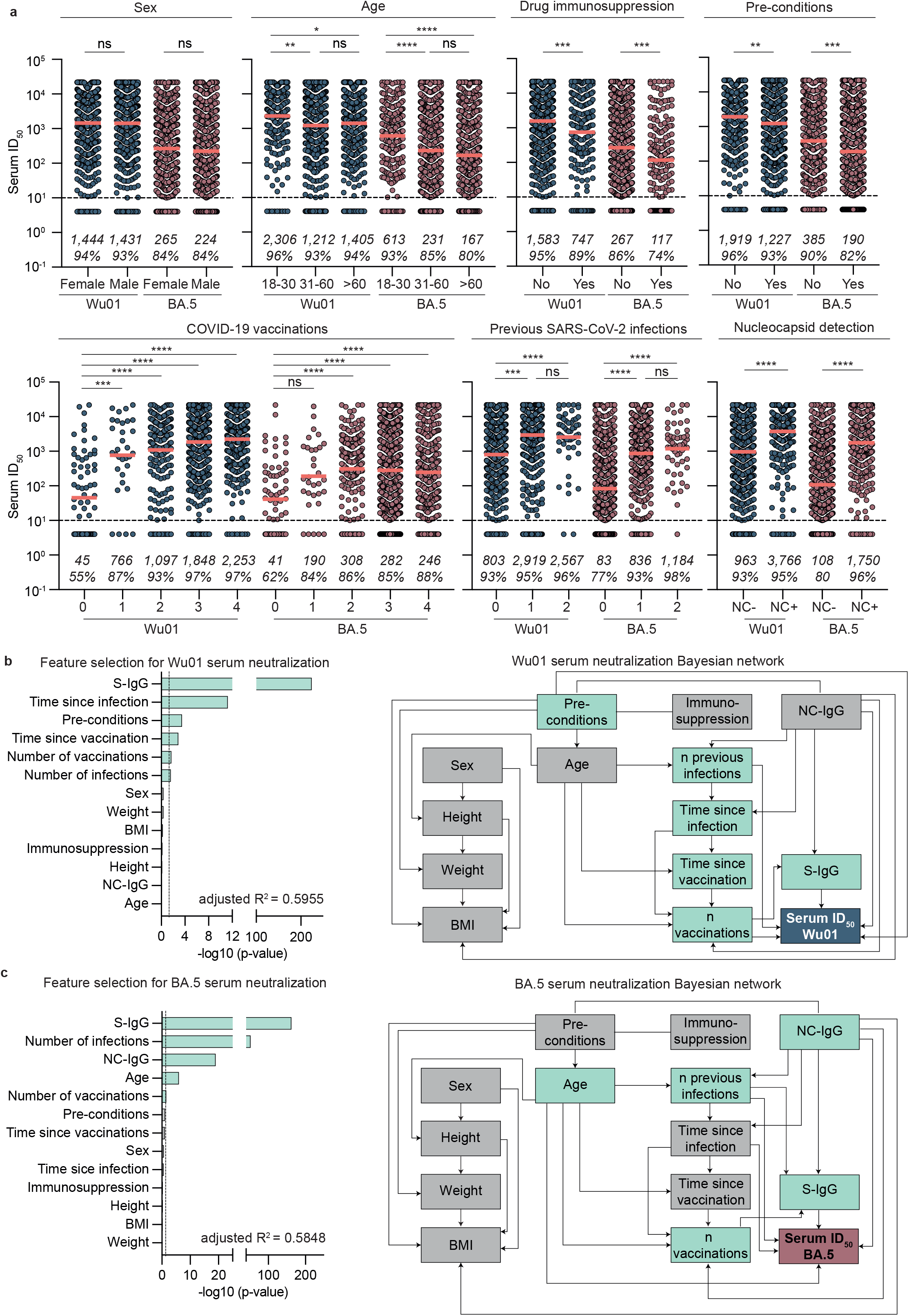
S-IgG levels and S-antigen contacts are most predictive for serum neutralization against Wu01 and BA.4/5. (a) Dot plots depicting serum neutralization ID_50_ values for Wu01 (blue) and BA.4/5 (red), subdivided based on sex, age, pre-conditions, drug immunosuppression, pre-conditions, number of vaccinations, number of infections, and NC-IgG detection. Dotted lines represent limit of detection (ID_50_ = 10). Geometric means are indicated by horizontal red lines and listed in each plot over total fractions of participants with detectable serum neutralization. Two-sided Kruskal-Wallis-tests, Dunn’s multiple comparisons tests and Mann-Whitney tests were performed for statistical analyses. Ns, *, **, ***, and **** represent p-values ≥ 0.05, < 0.05, ≤ 0.01, ≤ 0.001, and ≤ 0.0001, respectively. (b) Stepwise forward regression models for predicting serum neutralization (ID_50_) using continuous features (age, height, weight, body mass index (BMI), number of infections, number of vaccinations, time since infection, time since vaccination, S-IgG) and categorical features (sex, pre-conditions, NC-IgG and immunosuppression). Features with p < 0.05 in the multivariable regression model are highlighted in green. (c) Bayesian networks of the features predicting serum neutralization (ID_50_). The graph connects the features, which are predictive of each other with Serum neutralization as sink. Features with p < 0.05 in the multivariable regression model (B) are highlighted in green.

For serum neutralization against Wu01, multivariable regression (adjusted R^2^ = 0.5955) showed S-IgG levels as most predictive (p < 2×10^−16^), followed by time since infection (p = 0.012), pre-conditions (p = 0.0003), time since vaccination (p = 0.004), number of vaccinations (p = 0.003), and number of infections (p = 0.017). Bayesian network analysis revealed that S-IgG serostatus, NC-IgG serostatus, number of previous infections and vaccinations, and pre-conditions were directly predictive for serum neutralization against Wu01 (**Fig. 4b**). For serum neutralization against BA.4/5, the model (adjusted R^2^ = 0.5848) showed S-IgG levels as most predictive (p < 2×10^−16^), followed by number of previous infections (p < 2×10^−16^), NC-IgG serostatus (p < 2×10^−16^), age (p = 0.0001), and number of received vaccinations (p = 0.076). Bayesian network analysis revealed that S-IgG serostatus, NC-IgG serostatus, age, number of previous infections, and time since infection were directly predictive for serum neutralization against BA.4/5 (**Fig. 4c**).

We concluded that SARS-CoV-2 immune response is complex and determined by several factors. However, previous S-antigen contacts, that is, vaccinations and/or infections, substantially contribute to SARS-CoV-2 humoral immunity.

### Impaired serum neutralization activity against Omicron sub-lineage BQ.1.1

After completion of sample collection, a rapid spread of the BA.4/5-sublineage BQ.1.1 exhibiting three additional mutations in the Spike-protein in comparison to BA.4/5 (R346T, K444T, and N460K) could be observed (**Fig. 5a and b**). To determine serum neutralizing activity against BQ.1.1, we draw a proportionate stratified random sub-sample of 423 out of all 1,411 participants (29.9%) to be additionally tested in pseudovirus neutralization assay against BQ.1.1. Strata were defined by sex and 10-years age categories (**Supplementary Fig. 4a**). There were no significant differences in age and sex distributions (**Supplementary Fig. 4b**) as well as in S-IgG levels or neutralizing activity against Wu01 and BA.4/5 (**Supplementary Fig. 5**) between the sub-sample and the entire study population, confirming representativeness of the sub-sample for the entire study sample. While neutralizing activity against Wu01 and BA.4/5 was detectable in 93.9% and 84.9% of the participants, only 73.8% of the studied participants presented activity against the Omicron sub-lineage BQ.1.1. Geometric mean ID_50_s were 1,302, 231, and 55 (Friedmann test: p < 0.0001; Dunn’s multiple comparisons tests: p < 0.0001, p < 0.0001, p < 0.0001) for the respective variants. Overall, neutralizing activity against BQ.1.1 was reduced 23.6-fold and 4.2-fold in comparison to neutralizing activity against Wu01 and BA.4/5, respectively (**Fig. 5c**). The subsequent correlation of S-IgG values of all participants with detectable S-IgG against ID_50_ values of all participants with detectable serum neutralization (n = 294) against Wu01 (r_s_ = 0.64, p < 0.0001), BA.4/5 (r_s_ = 0.44, p < 0.0001), and BQ.1.1 (r_s_ = 0.48, p < 0.0001) revealed rather parallel fit lines and in comparison to Wu01 and a decrease of the intercepts for BA.4/5 and BQ.1.1, respectively (**Fig. 5d**, left panel). For S-IgG levels of 10-100 BAU/ml in 85.7%, and for S-IgG levels of 100-1,000 BAU/ml in 55.6% of the participants, no serum neutralization against BQ.1.1 could be detected (**Fig. 5d**, right panel). Spearman correlations between ID_50_ values of Wu01 versus BA.4/5 (r_s_ = 0.75), Wu01 versus BQ.1.1 (r_s_ = 0.67), and BA.4/5 versus BQ.1.1 (r_s_ = 0.78) revealed a stronger correlation between Wu01 and BA.4/5 serum neutralization than between Wu01 and BQ.1.1 serum neutralization, reflecting the improved immune escape of BQ.1.1 (**Fig. 5e**).

**Fig. 5:**
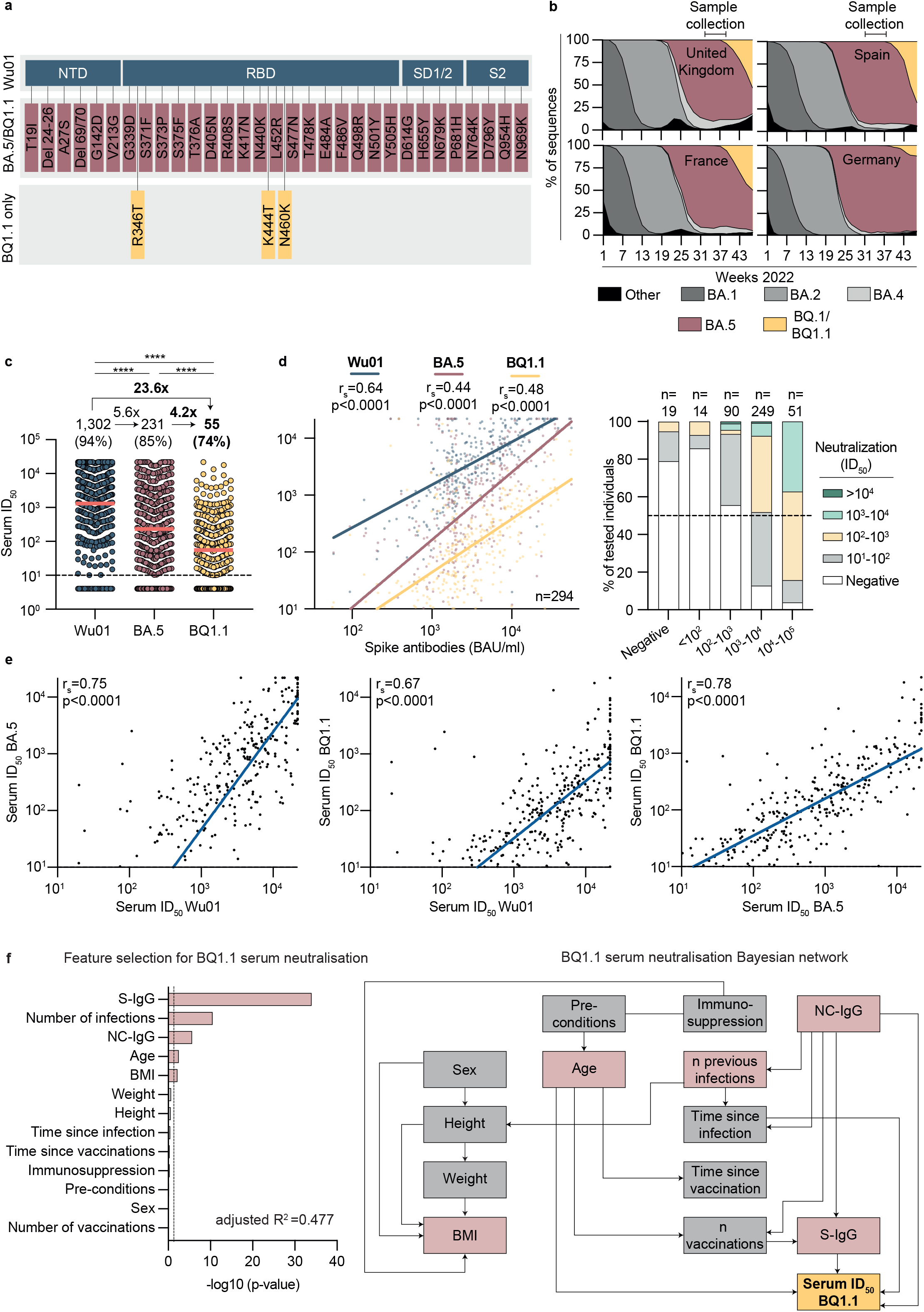
Impaired serum neutralization activity against Omicron sub-lineage BQ.1.1. (a) Spike amino acid changes in BQ.1.1 relative to BA.4/5 and Wu01. (b) Variant proportions in United Kingdom, Spain, France, and Germany extrapolated from bi-weekly Our world in Data dashboard and weekly reports of the Robert Koch institute (accessed on November 25, 2022). NTD, N-terminal domain; RBD, receptor-binding domain; S, Spike. (c) Dot plot depicting serum neutralization ID_50_ values for Wu01 (blue), BA.4/5 (red), and BQ.1.1 (yellow). Dotted lines represent the limit of detection (ID_50_ = 10). Geometric means are indicated by horizontal red lines and listed in each plot over total fractions of participants with detectable serum neutralization. Arrows indicates x-fold decrease of serum neutralization. Two-sided Friedman test and Dunn’s multiple comparison tests were performed for statistical analyses. Asterisks represent p-values < 0.05. (d) Left: Spearman correlation of Spike IgG (BAU/ml) against serum neutralization (ID_50_) of Wu01, BA.4/5 and BQ.1.1 variants indicated by respective colors. Red, blue and yellow lines indicate fit lines computed with robust regression. Right: Serum neutralization (ID_50_) is categorized by indicated color and stratified by S-IgG (BAU/ml) for BQ.1.1 variant. Dotted line represents 50% of all participants in each stratum. N of each stratum is indicated on top of the graph. (e) Spearman correlations of Wu01 ID_50_ against BA.4/5 ID_50_ (left), Wu01 ID_50_ against BQ.1.1 ID_50_ (middle) and BA.4/5 ID_50_ against BQ.1.1 ID_50_ (right). Blue lines indicate fit lines computed with robust regression (f) Stepwise forward regression models for predicting BQ.1.1 serum neutralization (ID_50_) using continuous features (age, height, weight, body mass index (BMI), number of infections, number of vaccinations, time since infection, time since vaccination, S-IgG) and categorical features (sex, pre-conditions, NC-IgG and immunosuppression). Features with p<0.05 in the multivariable regression model are highlighted in red. (g) Bayesian networks of the features predicting BQ.1.1 serum neutralization (ID_50_). The graph connects the features, which are predictive of each other with serum neutralization as sink. Features with p<0.05 in the multivariable regression model (F) are highlighted in red.

Finally, we performed multivariable regression and Bayesian network analysis as described above to further explore and predict serum neutralization against BQ.1.1. The resulting model (adjusted R^2^ = 0.477) showed S-IgG levels as most predictive (p = 2×10^−16^), followed by number of previous infections (p = 6.47×10^−5^) and NC-IgG serostatus (p = 2.7×10^−6^), age (p = 0.001), and BMI (p = 0.007), while all other features were not statistically significant. Bayesian network analysis revealed that S-IgG, NC-IgG, time since infection, and age were directly predictive for serum neutralization against BQ.1.1 (**Fig. 5f**).

We concluded that serum neutralization of the participants is impaired against BQ.1.1. Furthermore, previous S-antigen contacts are most predictive for serum neutralizing activity against BQ.1.1, as observed for Wu01 and BA.4/5.

## Discussion

Given the substantial immune escape of SARS-CoV-2 variants, for the assessment of population immunity, completing S-IgG seroprevalence with serum neutralizing activity is essential^26,27^. This can help to assess the COVID-19 risk and inform on public health measures, such as vaccination strategies for Omicron-adapted vaccines, mask requirements, and further concepts of prevention. At the time of sample collection, there was an urgent need for assessing the COVID-19 risk of vulnerable groups in autumn and winter 2022/2023 in Germany. For that reason, we conducted a cross-sectional point-seroprevalence study in patients that received medical treatment in emergency departments because we expected a high prevalence of pre-conditions and risk factors for severe courses of COVID-19 in those patients.

Our results demonstrate a high fraction of participants (95%) with detectable S-IgG but only moderate adherence to current recommendations on COVID-19 vaccinations (67%)^25^. The fraction of participants that reported previous infections (45%) was high and our results show that previous infections significantly contribute to neutralizing activity against SARS-CoV-2, which is in line with other studies^8,28–30^. Importantly, no data on the clinical course of the infections were available in our study. Accordingly, we do not conclude that infections rather than vaccinations represent a possible strategy in the future for boosting immunity in risk groups. Furthermore, in contrast to vaccinations, timing and outcome of infections are not predictable.

Most strikingly, we showed a 23-fold decrease in serum neutralizing activity against BQ.1.1 in comparison to Wu01. This is in line with to date limited data on BQ.1.1 immune escape and highlights BQ.1.1 as one of the variants with the highest extent of immune escape that have been observed so far^13–17,22,23,31,32^. We could not differentiate the immune escape to infection-induced antibody response and/or antibodies induced by mono- or bivalent vaccinations. However, substantial neutralization resistance of BQ.1.1 after BA.5 infections was shown previously^22^ and bivalent vaccination was shown to elicit lower neutralizing activity against BQ.1.1 than against BA.5^21^.

As shown in our multivariable regression model and Bayesian network analysis, S-IgG levels were most predictive for neutralization activity against BQ.1.1. However, we find it important to emphasize that 59.6% of the individuals with detectable S-IgG < 1000 BAU/ml showed no detectable neutralizing activity against BQ.1.1, highlighting the decreased accuracy of S-IgG detection for assessing neutralization in individuals with low S-IgG titers. This information is relevant for clinical routine testing of S-IgG and needs to be considered when assessing COVID-19 risk in patients.

To our best knowledge, this study is one of the largest that assesses immune evasion of BA.4/5 and BQ.1.1 in a real-life setting. Furthermore, the comprehensiveness of data on medical history, vaccinations, and previous infections of the participants contribute to a reliable assessment of humoral immunity of vulnerable persons before the worldwide predominance of BQ.1.1. However, our study population is not a representative sample of the general population and by that, external validity of S-IgG seroprevalence is limited. Nevertheless, we expect that the observed impaired serum neutralization against BQ.1.1 can be generalized, as it was controlled for several features including age, pre-conditions, drug immunosuppression and number of previous S-antigen contacts.

In summary, we determined a high S-IgG seroprevalence, only a moderate compliance with vaccination recommendations and subsequently a broad range of serum neutralizing activity against BQ.1.1. By that, the observed substantial fraction of persons without detectable neutralizing activity mirrors the consequences of the interplay between immune escape and non-compliance with vaccination recommendations. We conclude that improvement of vaccine-uptake for all eligible individuals is critical, predictable, and feasible for reducing the COVID-19 risk in upcoming waves of BQ.1.1 infections.

## Methods

### Ethical considerations

All samples and data were obtained under protocols approved by the ethics committees of the Medical Faculty of the University of Cologne (22_1262), of the Medical Faculty of the University of Bonn (314/22), of the Medical Faculty of the University of Düsseldorf (2022-2072), of the Medical Faculty of the University of Essen (22-10838-BO), and of the Medical Faculty of the University of Münster (2022-490-b-S). All participants provided written informed consent. This study was registered as clinical trial (DRKS00029414).

### Study design

Recruitment of participants and sample collection was conducted at five study sites in North Rhine-Westphalia, Germany (University Hospital of Cologne, University Hospital of Düsseldorf, University Hospital of Essen, University Hospital of Bonn, and University Hospital of Münster). Participation was offered to patients receiving medical treatment in emergency departments at one of the five study sites between August 8, 2022 and September 19, 2022. The study personnel recruited 1,411 participants in cooperation with the emergency department personnel. Patients were required to meet the following eligibility criteria in order to be enrolled as participants: i) only individuals aged ≥18 years were eligible, ii) participants had to be patients at the emergency department at one of the five study sites, iii) patients had to be able to consent to participate, iv) the ability to consent was furthermore checked by the study personnel with a special focus on the medication, pain and the exceptional emotional situation of the patients, and v) according to the assessment of the study personnel, participation in the study should not be a significant additional burden for the patients. After verification of eligibility criteria and obtaining written informed consent by the study physicians, during blood collection, which was part of the standard medical treatment at the emergency departments, up to additional 20 mL of blood was drawn from the participants to be analyzed in this study. Information on epidemiological data (age, sex, nationality, place of residence) and clinical data (body-length and weight, pre-existing conditions, immunosuppressive medication) as well as information on COVID-19 vaccination status (number of shots received, received vaccine type, date of vaccination), and past infections with SARS-CoV-2 (date of positive RT-qPCRs and/or rapid antigen detection tests) was verbally requested by the study personnel or extracted from the medical records of the participants. Samples and data collected at one study site were pseudonymized at that site and analyzed centrally at the study site in Cologne. In addition, serum samples obtained at each study site were analyzed at that site for assessing assay validity as described further in Methods Details.

### Processing of serum samples

Samples were transported at 4°C, processed within 48 hours after collection by centrifugation and stored at −80°C till use.

### Serological assays

Anti-SARS-CoV-2 antibodies were detected using commercial assays that use either SARS-CoV-2 Spike protein or SARS-CoV-2 Nucleocapsid antigens. All assays were used as per manufacturer’s recommendations.

S-IgG were measured using DiaSorin’s LIAISON® SARS-CoV-2 TrimericS chemiluminescence immunoassay as described previously^33^ with the following cut-off values: negative < 33.8 BAU/ml and positive ≥ 33.8 BAU/ml. Positive percent agreement (PPA) and negative percent agreement (NPA) with i) Anti-SARS-CoV-2 QuantiVac IgG BAU (Euroimmun) (n = 502, measured at the study site in Düsseldorf) were 99.78% and 83.33%, with ii) DiaSorin’s LIAISON® SARS-CoV-2 TrimericS chemiluminescence immunoassay (n = 185, measured at the study site in Essen) were 100% and 100%, with iii) Abbott’s anti-SARS-CoV-2 IgG Quant II chemiluminescence microparticle assay (Alinity i) (n = 133, measured at the study site in Bonn) were 100% and 66.66% (n = 3), and with iv) Abbott’s anti-SARS-CoV-2 IgG Quant II chemiluminescence microparticle assay (Alinity i) (n = 208, measured at the study site in Münster) were 99.5% and 100%. For graphical representation and statistical evaluation of serum samples, in Figures 2, 3, 5, Figure S5, samples that did not achieve IgG levels ≥33.8 BAU/ml were imputed to 4.81 BAU/ml (lower limit of quantification).

NC-IgG were measured using the Euroimmun anti-SARS-CoV-2-NCP-ELISA. Serum samples were tested on the automated system Euroimmun Analyzer I according to manufacturer’s recommendations. S/CO values were interpreted as positive (S/CO ≥1.1), borderline (S/CO ≥ 0.8 - < 1.1), and negative (S/CO < 0.8). PPA and NPA with i) Abbott’s Architect SARS-CoV-2 IgG assay (n = 502, measured at the study site in Düsseldorf) were 97.6% and 94.85%, with ii) Abbott’s Architect SARS-CoV-2 IgG assay (n = 185, measured at the study site in Essen) were 100% and 95.95%, with iii) Roche’s Elecsys®-Assay (n = 133, measured at the study site in Bonn) were 56.86% and 100%, and with iv) Abbott’s Architect SARS-CoV-2 IgG (n = 208, measured at the study site in Münster) were 83.05% and 98.92%.

### Cell lines

HEK293T cells and 293T-ACE2 cells were maintained in DMEM (Gibco) containing 10% FBS, 1% Penicillin-Streptomycin, 1mM L-Glutamine and 1mM Sodium pyruvate. Cells were grown in T75 flasks (Sarstedt) at 37°C and 5% CO_2_.

### Cloning of SARS-CoV-2 Omicron BA.**4/5 and BQ.1.1 spike constructs**

Cloning of Wu01- and BA.4/5 spike protein expression plasmids was previously described^13,34^. Compared with the Wu01 strain spike protein amino acid sequence, for the Omicron BA.4/5 strain, the following changes were included in the plasmid: T19I, Δ24-26, A27S, D69-70 G142D, V213G, G339D, S371F, S373P, S375F, T376A, D405N, R408S, K417N, N440K, L452R S477N, T478K, E484A, F486V Q498R, N501Y, Y505H, D614G, H655Y, N679K, P681H, N764K, D796Y, Q954H, and N969K mutations. For the BQ.1.1 spike protein expression plasmid, a gene fragment (Thermo Fisher) encompassing the additional R346T, K444T, and N460K mutations was cloned into the BA.4/5 spike protein expression plasmid using the NEB HiFi DNA Assembly Kit (New England Biolabs). All spike protein expression plasmids incorporate a C-terminal deletion of 21 cytoplasmic amino acids that results in increased pseudovirus titers. Sanger sequencing was used for verification of the spike sequence.

### Pseudovirus neutralization assays

For pseudovirus particle production, HEK293T cells were co-transfected with plasmids encoding for the SARS-CoV-2 spike protein, HIV-1 Tat, HIV-1 Gag/Pol, HIV-1 Rev, and luciferase followed by an internal ribosome entry site (IRES) and ZsGreen^35^. FuGENE 6 Transfection Reagent (Promega) was used for transfection. Virus culture medium was harvested 48-72h after transfection and stored at -80°C. Harvested virus was titrated. To this end, 293T-ACE2 cells^35^ were infected and incubated 48 h at 37°C and 5% CO_2_ prior to luciferase activity assessment. Activity was determined using a microplate reader (Berthold) after addition of luciferin/lysis buffer (10 mM MgCl2, 0.3 mM ATM, 0.5 mM Coenzyme A, 17 mM IGEPAL (all Sigma-Aldrich), and 1 mM D-Luciferin (GoldBio) in Tris-HCL). After heat inactivation (56°C at 45 min), serum samples were serially diluted (1:3), starting with a 1:10 dilution. Before the addition of 293T-ACE2 cells, dilutions of serum samples were co-incubated with pseudovirus supernatants for 1h at 37°C. All samples were tested in single dilution series. Using the reagents described above, luciferase activity was determined after 48h incubation at 37°C and 5% CO_2_. To quantify neutralization activity, after subtraction of background RLUs of non-infected cells, the 50% inhibitory dose (ID_50_) was determined. ID_50_ was defined as the serum dilution which resulted in a 50% reduction in RLUs in comparison with the untreated virus control cells. GraphPad Prism 9 was used for calculation of ID_50_ which was plotted as dose response curve. A SARS-CoV-2 neutralizing monoclonal antibody was used as run control. Assay specificity was described before^36^. Average inter-assay coefficient was determined as 18.09%, by testing 1,546 serum samples in duplicates on different plates and different days. For graphical representation and statistical evaluation of serum samples in Figures 3, 4, 5 and Figure S5 samples that did not achieve 50% inhibition at the lowest tested dilution of 10 (lower limit of quantification, LLOQ) were imputed to ID_50_ = 4 and serum samples with ID_50_s >21,870 (upper limit of quantification) were imputed to ID_50_ = 21,871.

### SARS-CoV-2 variant frequencies

SARS-CoV-2 variant proportions were extrapolated from bi-weekly Our world in Data dashboard (http://ourworldindata.org, accessed on November 25, 2022) and weekly reports of the Robert Koch Institute.

### Quantification and statistical analysis

We used a stepwise forward regression to select features in a linear regression model. We distinguished between continuous features (age, height, weight, BMI, S-IgG, serum ID_50_ Wu01, serum ID_50_ BA.4/5, serum ID_50_ BQ.1.1, number of previous infections, number of vaccinations, time since infection and time since vaccination) and categorical features (sex, preconditions, NC-IgG and drug immunosuppression) (**Figures 2d, 4b, 4c, 5f)**. We removed all patient samples with no data for at least one feature, either missing or not specified. Our final data set had a size of 1,209, 1,209, 1,207 and 352 for S-IgG, serum ID_50_ Wu01, serum ID_50_ BA.4/5, and serum ID_50_ BQ.1.1, respectively. We transformed the features with a skewed distribution (S-IgG, Serum ID_50_, and time since infection) by adding 1 and taking the log with base 10. We used stepwise forward regression for feature selection. We started with the base model, only including the intercept and iteratively added the feature that increased the model fit the most. We tested the increase of the model fit with a likelihood-ratio test (R package lmtest). We did not include any Serum ID_50_ features as predictors. We standardized the continuous features in our final model with mean zero and standard deviation 1 to allow for the comparison of the coefficients. For the next analysis, the larger data sets shrunk to 1,181 participants, because we included all features into one model, which led to more missing data in several participants. We created 1,000 different datasets by sampling 1,181 respectively 352 patients with replacement from the original participant dataset. We used the score-based hill-climbing algorithm (R package bnlearn) to compute a Bayesian network for each dataset (**Figures 2e, 4b, 4c, 5f**). We restricted the response from having outgoing edges to preserve the directionality of the regression. We computed the edge fractions from the 1,000 Bayesian networks and defined a consensus network by including edges with a fraction of 0.5, i.e., 500 or more appearances.

Testing for statistical significance of differences in sex distribution between the study population and the study sample was performed with the two-sided Fisher’s exact test. Testing for statistical significance of differences in S-IgG levels, NC-IgG S/CO values or serum neutralization titers against Wu01, BA.4/5 and BQ.1.1 was performed with the two-sided Kruskal-Wallis test, Friedman test, Mann-Whitney U test or Wilcoxon matched pairs signed rank test, using Prism 9.0 (GraphPad). Dunn’s multiple comparisons tests were performed as post-hoc tests (GraphPad). Spearman’s rank correlation coefficients (R_s_) were determined using Prism 9.0 (GraphPad). Non-linear regression (robust regression) was calculated using Prism 9.0 (GraphPad). Statistical significance was defined as p < 0.05. Details are additionally provided in the Figure legends.

### Additional software

Maps of Germany and North Rhine-Westphalia were designed with the iMapU tool provided by iExcelU.

## Data Availability

All data produced in the present study are available upon reasonable request to the authors

## Data availability

Raw data reported in this paper will be shared by the lead contact upon reasonable request.

## Code availability

All original code has been deposited online^37^ and is publicly available as of the date of publication.

## Acknowledgements

We are grateful to all study participants for their dedication to our research. We thank all members of all involved institutes. In particular, we thank Andrea Klöckner, Christian Overath, Anna Lucia Petrucci, Flaurant Ramadani, Claudia Reddig, Jasmin Lange, and Monika Eschbach-Bludau for processing of serum samples and sample logistics.We thank Kanika Vanshylla for the establishing of the pseudovirus neutralization assay. We thank Thorsten Noack-Schönborn for providing data on emergency department patient characteristics. We thank Carsten Tschirner (IExcelU) for supporting data visualization and all laboratories providing SARS-CoV-2 sequencing information to GISAID and subsequently Our world in Data for facilitating variant distribution analyses.

## Author contributions

Conceptualization, F.D., F.K., J.T., S.L., H.S., C.E., M.P., B.S., U.D.; J.Kü.; methodology, F.D., M.P., F.K.; investigation, F.D., M.P., F.K., E.A. B.M., V.D.C, V.B., H.G., F.T., M.B., M.M., M.L., M.A., M.T.H., W.H., M.M.M., P.K., J.Kü., M.S., G.O., J.K., C.E., C.K., D.E., I.G., M.K., W.O.M.-P., J.R., A.B., M.B., R.K.M., M.W., U.V.F., E.R.; resources, F.K., J.T., H.S. U.D., S.L.; formal analysis, F.D., M.P., and F.K; visualization, F.D.; writing - original draft, F.D., M.P., and F.K.; writing - review & editing, all authors; supervision, F.K.; funding acquisition, F.K., J.T., H.S. U.D., and S.L.

## Additional Information

Supplementary Information is available for this paper.

Correspondence should be addressed to.

Funding was provided by the ministry for work, health, and social affairs of the state of North Rhine-Westphalia (CPS-1-1F).

## Supplementary information

**Supplementary Figure 1:**
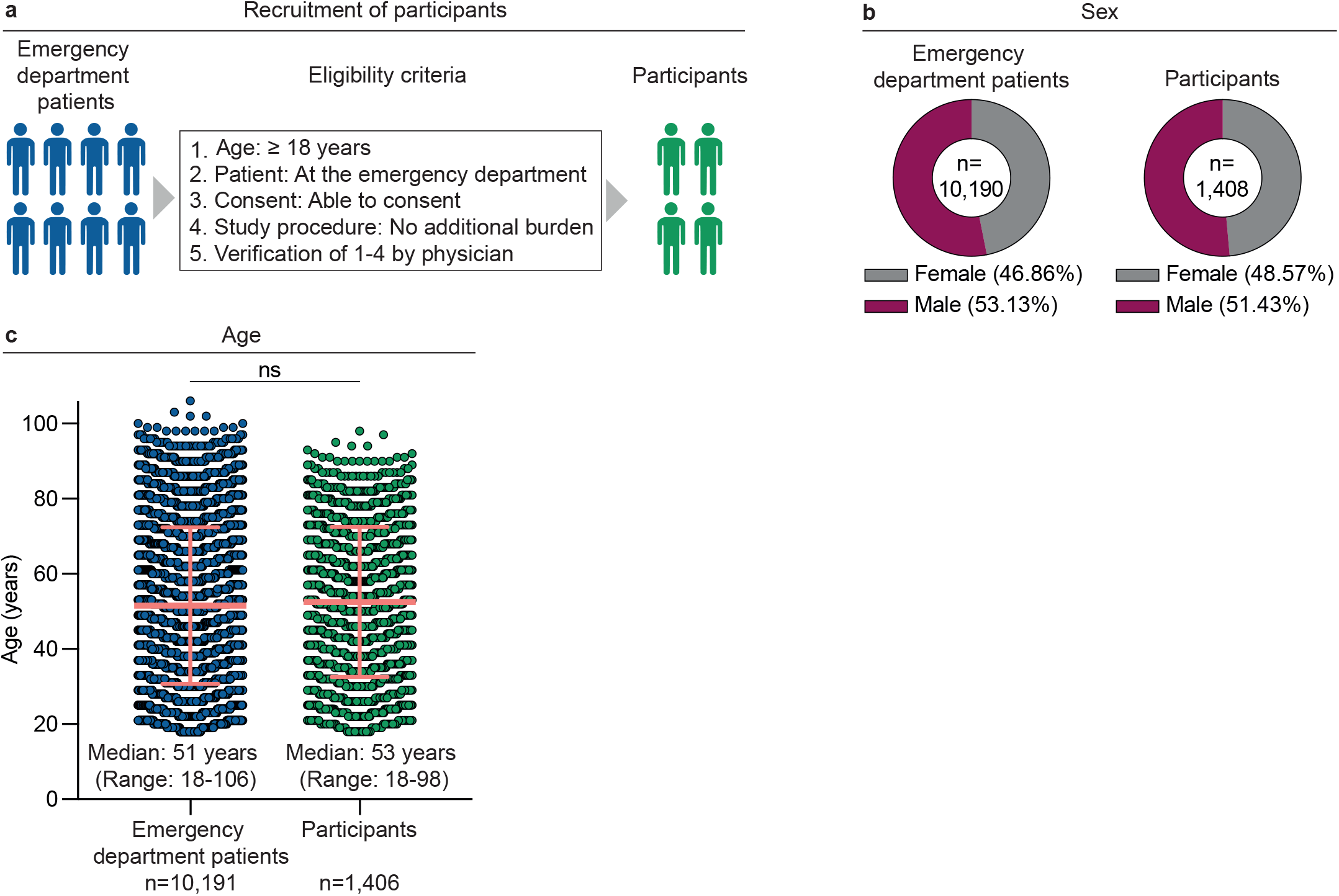
Recruitment of emergency department patients. (a) Illustration depicting eligibility criteria for participation. (b) Pie charts illustrating sex distribution of study population and sample. Sex of 1 patient and 3 participants was not available. Two-sided Fisher’s exact test (p=0.231) was performed for statistical analyses. (c) Dot plot illustrating age distribution of study population and sample. Age of 5 participants was not available. Median age is indicated by horizontal line, standard deviation is indicated by error bars. Mann-Whitney test was performed for statistical analyses. Ns indicates a p-value < 0.05.

**Supplementary Figure 2:**
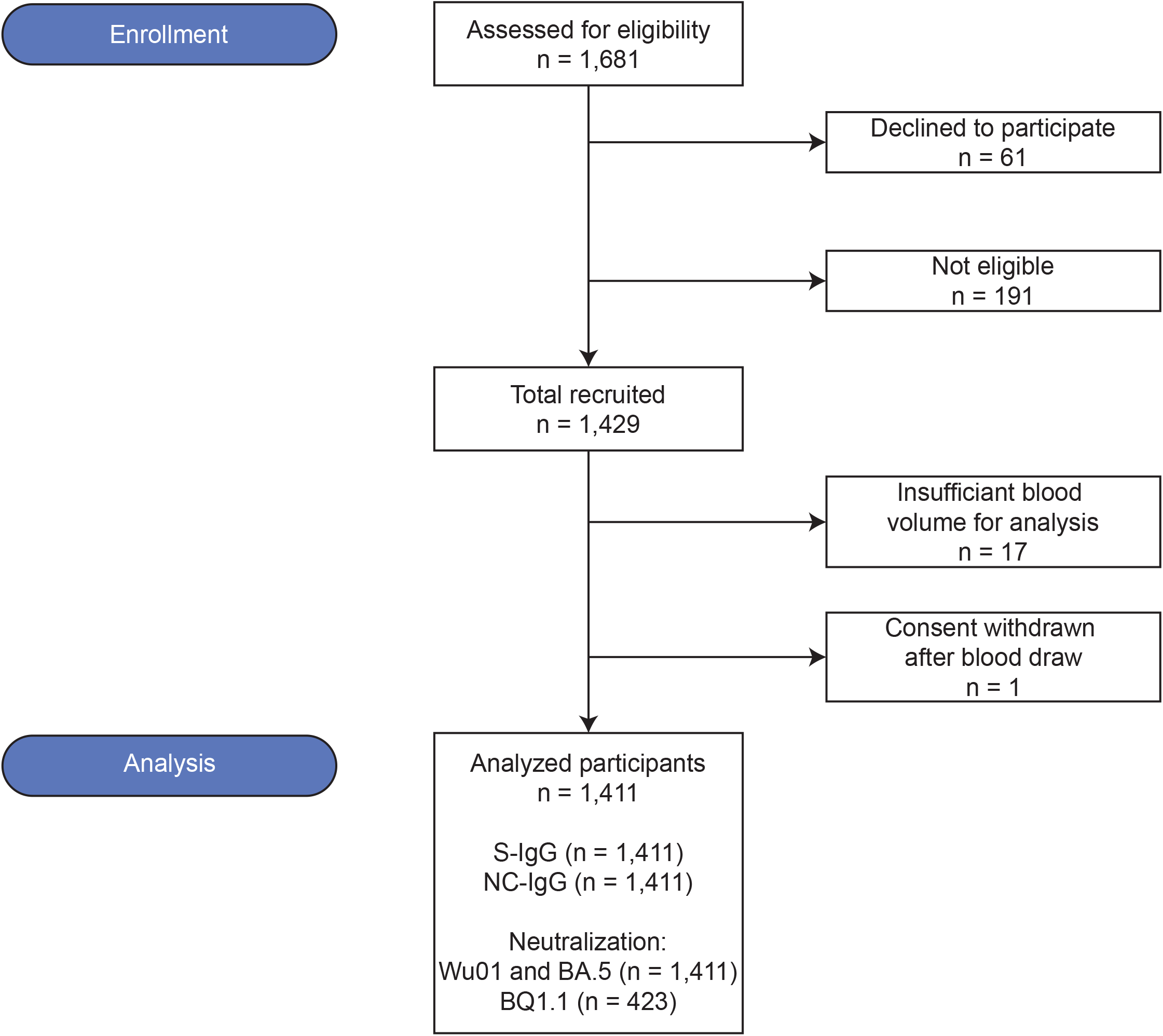
CONSORT flow diagram.

**Supplementary Figure 3:**
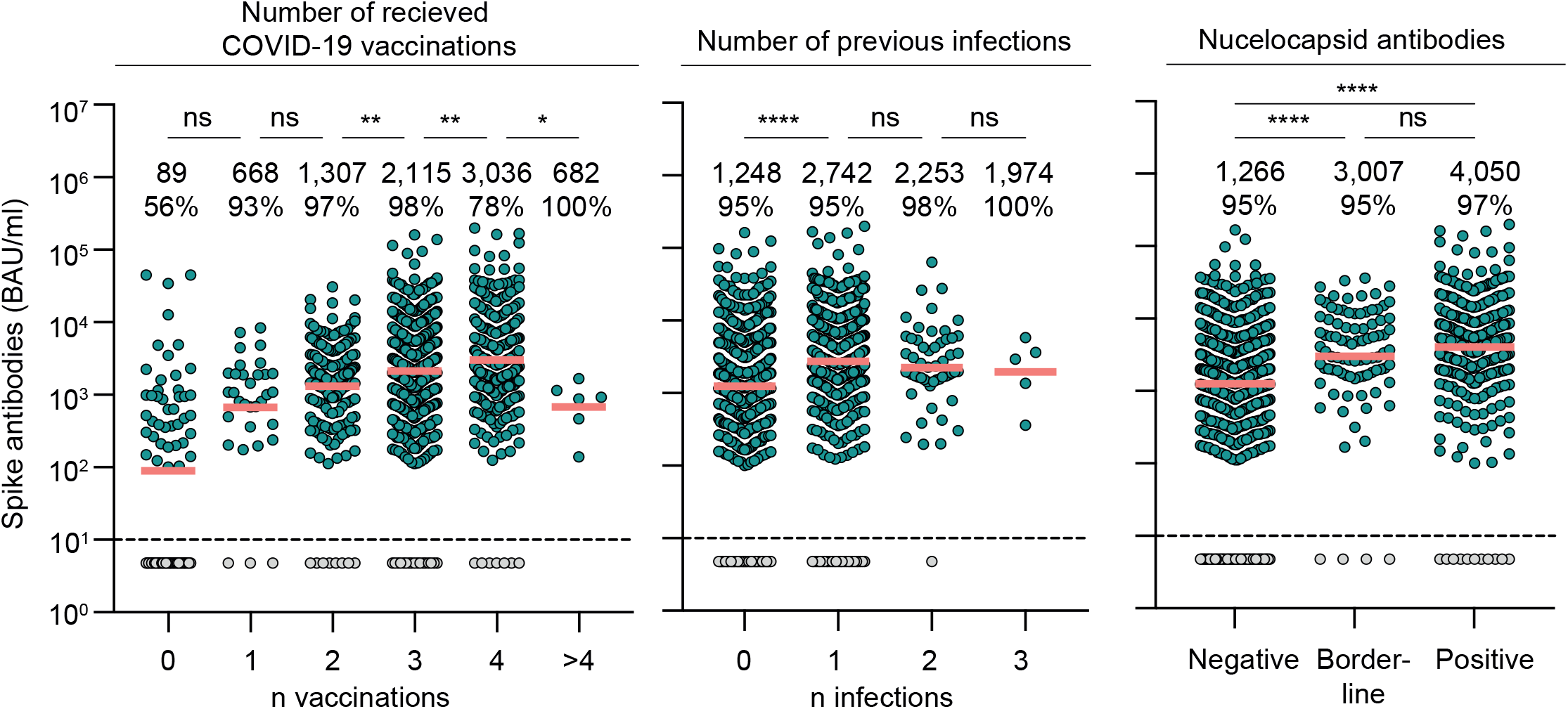
SARS-CoV-2 antibody prevalence in emergency room patients (additional features) Dot plots depicting S-IgG BAU/ml values, subdivided based on number of reported vaccinations, number of reported infections, and NC-IgG detection. Dotted lines represent the limit of detection (33.8 BAU/ml). Geometric means are indicated by horizontal red lines and listed in each plot over total fractions of participants with detectable Spike IgG. Two-sided Kruskal-Wallis-tests and Dunn’s multiple comparisons tests were performed for statistical analyses. Ns, *, **, ***, and **** represent p-values ≥ 0.05, < 0.05, ≤ 0.01, ≤ 0.001, and ≤ 0.0001, respectively.

**Supplementary Figure 4:**
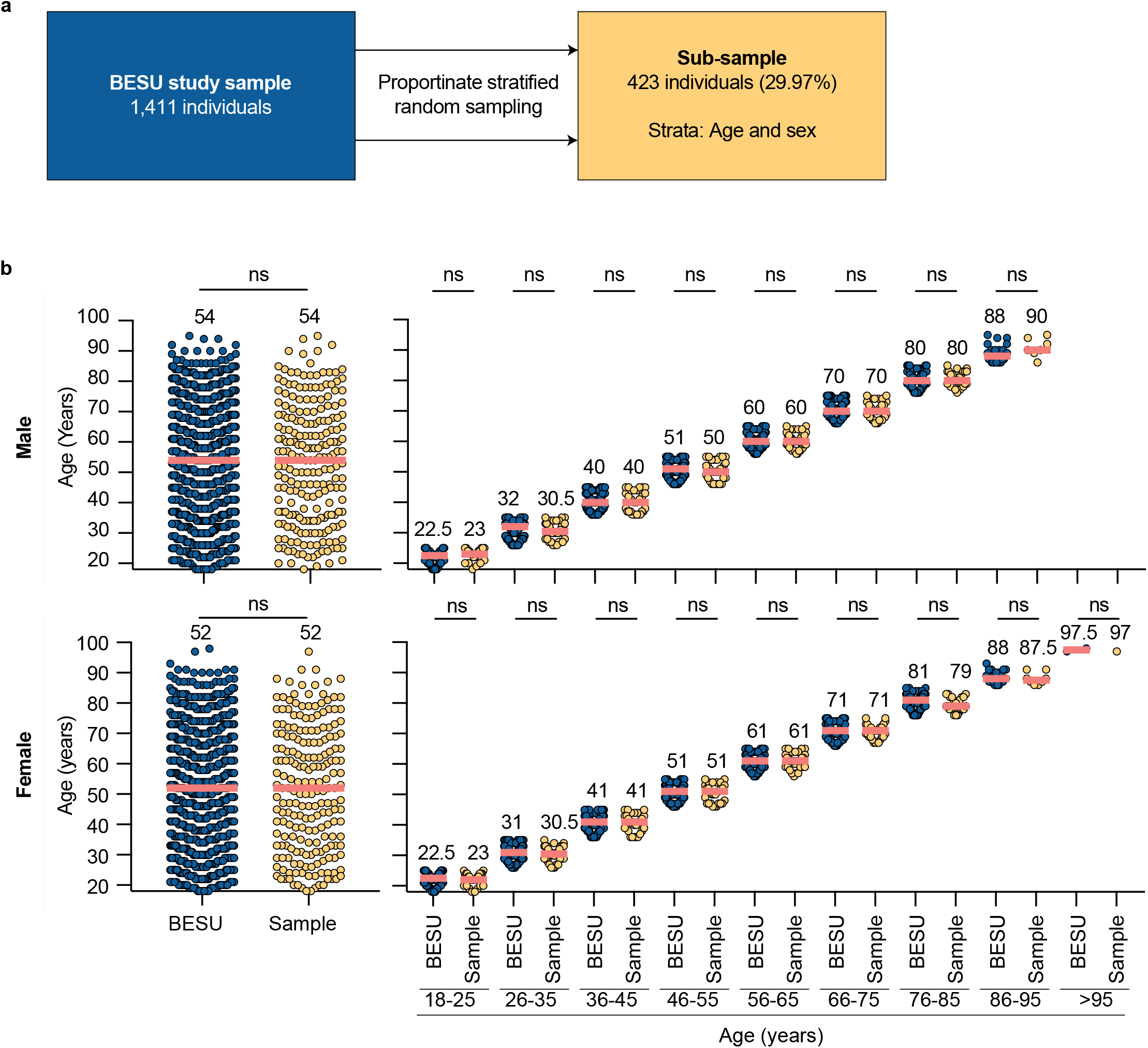
Sub-sampling for determination of BQ1.1 serum neutralization (Age and sex) (a) Illustration depicting statistical procedure for sub-sampling. Proportionate stratified random sampling was conducted to draw a representative sub-sample (n=423). (b) Dot plots depicting age distributions of all enrolled participants (BESU study sample) and the sub-sample, subdivided by sex and age-strata. Median ages are indicated by horizontal red lines and given above the dots. Two-sided Mann-Whitney tests and Kruskal-Wallis tests were performed for statistical analyses. Ns represents p-values > 0.05.

**Supplementary Figure 5:**
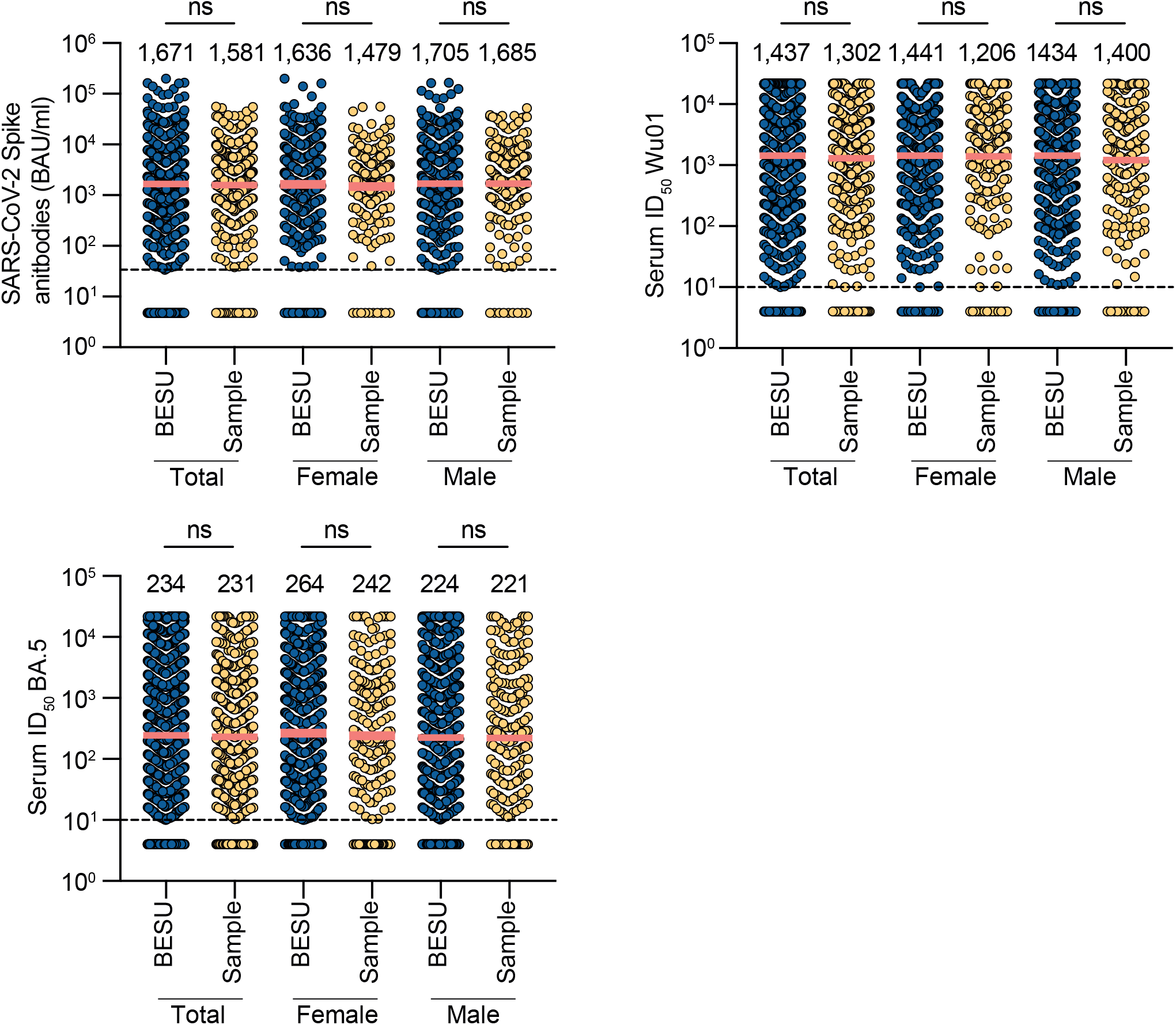
Sub-sampling for determination of BQ1.1 serum neutralization (S-IgG and ID_50_) Dot plots depicting S-IgG values (BAU/ml) and serum neutralization (ID50) of the entire study population and the sub-sample stratified by sex. Geometric means are indicated by horizontal red lines and given above the dots. Two-sided Mann-Whitney tests and Kruskal-Wallis tests were performed for statistical analyses. Ns represents p-values > 0.05.

**Supplementary Table 1:**
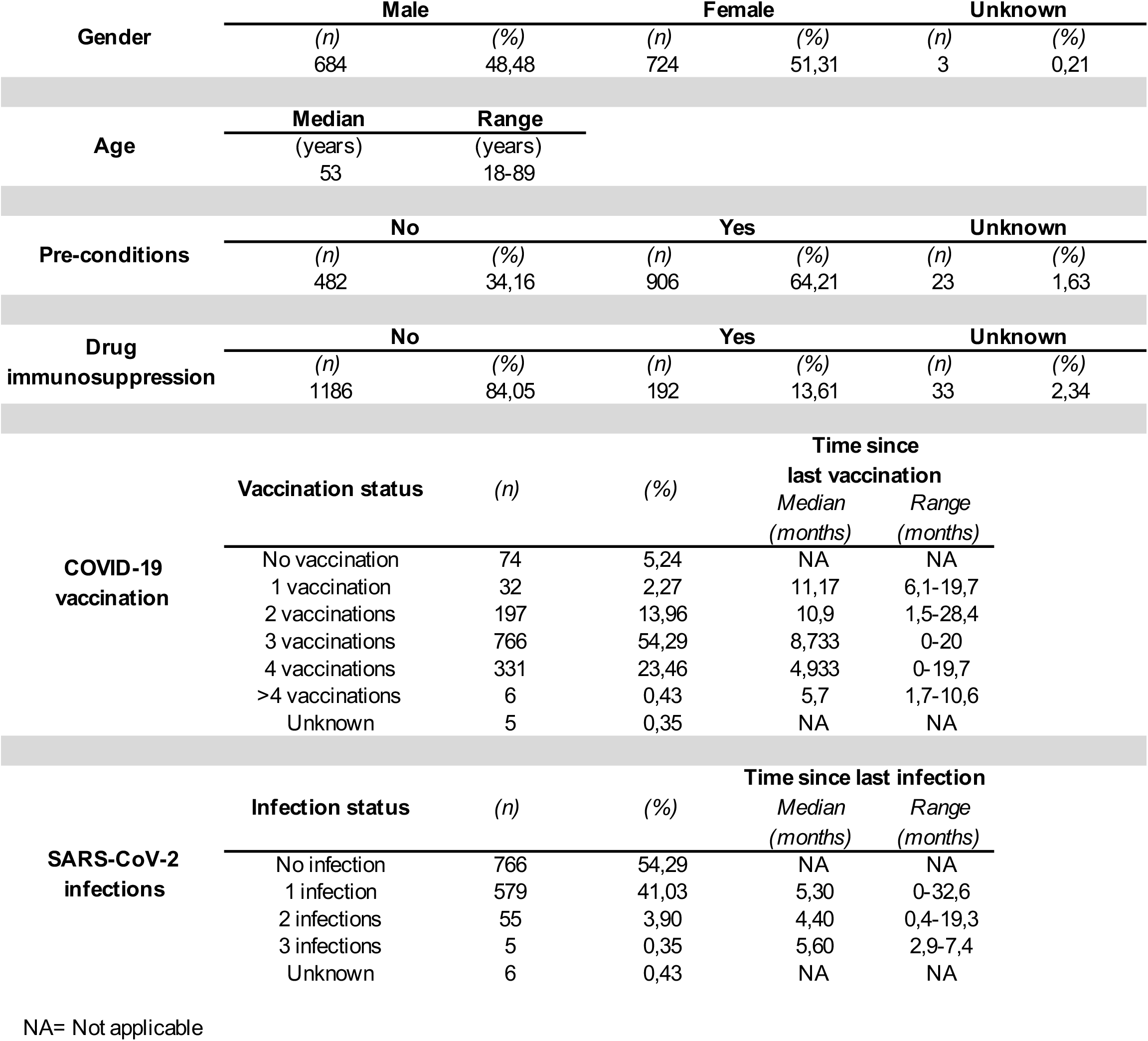
Study demographics, clinical information, and SARS-CoV-2 immune status.

## References

1. Lopez Bernal, J. et al. Effectiveness of Covid-19 Vaccines against the B.1.617.2 (Delta) Variant. N Engl J Med 385, 585–594 (2021).

2. Tseng, H. F. et al. Effectiveness of mRNA-1273 against SARS-CoV-2 Omicron and Delta variants. Nature Medicine 2022 28:5 28, 1063–1071 (2022).

3. Nyberg, T. et al. Comparative analysis of the risks of hospitalisation and death associated with SARS-CoV-2 omicron (B.1.1.529) and delta (B.1.617.2) variants in England: a cohort study. The Lancet 399, 1303–1312 (2022).

4. Tran, T. N. A. et al. SARS-CoV-2 Attack Rate and Population Immunity in Southern New England, March 2020 to May 2021. JAMA Netw Open 5, e2214171–e2214171 (2022).

5. Aziz, N. A. et al. Seroprevalence and correlates of SARS-CoV-2 neutralizing antibodies from a population-based study in Bonn, Germany. Nature Communications 2021 12:1 12, 1–10 (2021).

6. Stringhini, S. et al. Seroprevalence of anti-SARS-CoV-2 IgG antibodies in Geneva, Switzerland (SEROCoV-POP): a population-based study. The Lancet 396, 313–319 (2020).

7. Goldberg, Y. et al. Protection and Waning of Natural and Hybrid Immunity to SARS-CoV-2. New England Journal of Medicine 386, 2201–2212 (2022).

8. Suryawanshi, R. & Ott, M. SARS-CoV-2 hybrid immunity: silver bullet or silver lining? Nature Reviews Immunology 2022 22:10 22, 591–592 (2022).

9. Karim, S. S. A. & Karim, Q. A. Omicron SARS-CoV-2 variant: a new chapter in the COVID-19 pandemic. Lancet 6736, 19–21 (2021).

10. Brandal, L. T. et al. Outbreak caused by the SARS-CoV-2 Omicron variant in Norway, November to December 2021. Eurosurveillance 26, 1–5 (2021).

11. Pulliam, J. R. C. et al. Increased risk of SARS-CoV-2 reinfection associated with emergence of Omicron in South Africa. Science (1979) 376, (2022).

12. Tegally, H. et al. Emergence of SARS-CoV-2 Omicron lineages BA.4 and BA.5 in South Africa. Nature Medicine 2022 28:9 28, 1785–1790 (2022).

13. Gruell, H. et al. SARS-CoV-2 Omicron sublineages exhibit distinct antibody escape patterns. Cell Host Microbe 30, 1231-1241.e6 (2022).

14. Cao, Y. et al. BA.2.12.1, BA.4 and BA.5 escape antibodies elicited by Omicron infection. Nature 2022 608:7923 608, 593–602 (2022).

15. Tuekprakhon, A. et al. Antibody escape of SARS-CoV-2 Omicron BA.4 and BA.5 from vaccine and BA.1 serum. Cell 185, 2422-2433.e13 (2022).

16. Khan, K. et al. Omicron BA.4/BA.5 escape neutralizing immunity elicited by BA.1 infection. Nature Communications 2022 13:1 13, 1–7 (2022).

17. Hachmann, N. P. et al. Neutralization Escape by SARS-CoV-2 Omicron Subvariants BA.2.12.1, BA.4, and BA.5. New England Journal of Medicine 387, 86–88 (2022).

18. Mathieu, E. et al. Coronavirus Pandemic (COVID-19). (2020).

19. TAG-VE statement on Omicron sublineages BQ.1 and XBB. https://www.who.int/news/item/27-10-2022-tag-ve-statement-on-omicron-sublineages-bq.1-and-xbb.

20. Arora, P. et al. Omicron sublineage BQ.1.1 resistance to monoclonal antibodies. Lancet Infect Dis 0, (2022).

21. Zou, J. et al. Improved Neutralization of Omicron BA.4/5, BA.4.6, BA.2.75.2, BQ.1.1, and XBB.1 with Bivalent BA.4/5 Vaccine. doi:10.1101/2022.11.17.516898.

22. Qu, P. et al. Enhanced Neutralization Resistance of SARS-CoV-2 Omicron Subvariants BQ.1, BQ.1.1, BA.4.6, BF.7 and BA.2.75.2. Cell Host Microbe (2022) doi:10.1016/J.CHOM.2022.11.012.

23. Wang, Q. et al. Alarming antibody evasion properties of rising SARS-CoV-2 BQ and XBB subvariants. Cell (2022) doi:10.1016/J.CELL.2022.12.018.

24. Ito, J. et al. Convergent evolution of the SARS-CoV-2 Omicron subvariants leading to the emergence of BQ.1.1 variant. bioRxiv 11, 2022.12.05.519085 (2022).

25. Matysiak-Klose, D. et al. Empfehlung und wissenschaftliche Begründung der STIKO zur Grundimmunisierung von Personen im Alter von 12 – 17 Jahren mit dem COVID-19-Impfstoff Nuvaxovid von Novavax. Epid Bull 52–56 (2022).

26. Ng, D. L. et al. SARS-CoV-2 seroprevalence and neutralizing activity in donor and patient blood. Nature Communications 2020 11:1 11, 1–7 (2020).

27. Crawford, K. H. D. et al. Protocol and Reagents for Pseudotyping Lentiviral Particles with SARS-CoV-2 Spike Protein for Neutralization Assays. Viruses 2020, Vol. 12, Page 513 12, 513 (2020).

28. Ntziora, F. et al. Protection of vaccination versus hybrid immunity against infection with COVID-19 Omicron variants among Health-Care Workers. Vaccine 40, (2022).

29. Goldberg, Y. et al. Protection and Waning of Natural and Hybrid Immunity to SARS-CoV-2. New England Journal of Medicine 386, 2201–2212 (2022).

30. Gazit, S. et al. Severe Acute Respiratory Syndrome Coronavirus 2 (SARS-CoV-2) Naturally Acquired Immunity versus Vaccine-induced Immunity, Reinfections versus Breakthrough Infections: A Retrospective Cohort Study. Clinical Infectious Diseases 75, e545–e551 (2022).

31. Gruell, H. et al. Antibody-mediated neutralization of SARS-CoV-2. Immunity 55, 925–944 (2022).

32. Cao, Y. et al. Imprinted SARS-CoV-2 humoral immunity induces convergent Omicron RBD evolution. bioRxiv 2022.09.15.507787 (2022) doi:10.1101/2022.09.15.507787.

33. Affeldt, P. et al. Immune Response to Third and Fourth COVID-19 Vaccination in Hemodialysis Patients and Kidney Transplant Recipients. Viruses 2022, Vol. 14, Page 2646 14, 2646 (2022).

34. Vanshylla, K. et al. Discovery of ultrapotent broadly neutralizing antibodies from SARS-CoV-2 elite neutralizers. Cell Host Microbe 30, 69-82.e10 (2022).

35. Crawford, K. H. D. et al. Protocol and Reagents for Pseudotyping Lentiviral Particles with SARS-CoV-2 Spike Protein for Neutralization Assays. Viruses 12, (2020).

36. Vanshylla, K. et al. Kinetics and correlates of the neutralizing antibody response to SARS-CoV-2 infection in humans. Cell Host Microbe 29, 917-929.e4 (2021).

37. Pirkl, M. Impaired humoral immunity to BQ.1.1 in convalescent and vaccinated patients. Preprint at https://doi.org/https://10.5281/zenodo.7466593 (2022).

